# Anticipating the risk and spatial spread of measles in populations with high MMR uptake: using school-household networks to understand the 2013 - 2014 outbreak in the Netherlands

**DOI:** 10.1101/2024.02.20.24302866

**Authors:** James D Munday, Katherine E Atkins, Don Klinkenberg, Marc Meurs, Erik Fleur, Susan Hahne, Jacco Wallinga, Albert Jan van Hoek

## Abstract

Measles outbreaks are still routine, even in countries where vaccination coverage exceeds the guideline of 95%. Therefore, achieving ambitions for measles eradication will require understanding how unvaccinated children interact with others who are unvaccinated. Here we propose a novel framework for modelling measles transmission to better understand outbreaks in high uptake situations.

The high importance of school- and home-based transmission to overall outbreak dynamics is well established. Making use of this, we created a network of all primary and secondary schools in the Netherlands based on the total number of household pairs between each school. A household pair are siblings from the same household who attend a different school. We parameterised the network with individual level administrative household data provided by the Dutch Ministry for Education and estimates of school level uptake of the Mumps, Measles and Rubella (MMR) vaccine. We analyse the network to establish the relative strength of contact between schools. We simulated measles outbreaks on the network and evaluated the model against empirical measles data per postcode-area from a large outbreak in 2013 (2766 cases), comparing the model to alternative models that do not account for specific network structure or school-level vaccine uptake.

Our network analysis shows that schools associated with low vaccine uptake are highly connected, particularly Orthodox-Protestant schools (Coleman Homophily Index = 0.63). Simulations on the Network were able to reproduce the observed size and spatial distribution of the historic outbreak much more clearly than the alternative models, with a case weighted Receiver Operating Condition sensitivity of 0.94 for the data-driven network model and 0.38 and 0.23 for the alternative models. Further, we establish that variation in local network properties result in clear differences in final size of outbreaks seeded in orthodox-protestant-affiliated and other schools with low MMR coverage.

Our framework indicates that clustering of unvaccinated children in primary schools connected by unvaccinated children in related secondary schools lead to large, connected clusters of unvaccinated children. Using our approach, we could explain historical outbreaks on a spatial level. Our framework could be further developed to aid future outbreak response.

## Introduction

The World Health Organization has outlined ambitious goals for eliminating measles [1]. Despite the effectiveness of the Measles, Mumps, and Rubella (MMR) vaccine, estimates suggest that about 95 percent of the population needs to be vaccinated to achieve herd immunity, which is a level that prevents widespread disease transmission [2–5]. However, many countries are facing challenges due to increasing numbers of unvaccinated children. This rise is driven by parental hesitancy towards vaccines and reduced MMR vaccination rates during and after the COVID-19 pandemic [6,7].

Interestingly, even in countries where more than 95 percent of people have received at least one dose of the MMR vaccine, measles outbreaks still occur. This is because the traditional concept of herd immunity assumes that vaccines are distributed evenly throughout the population [8]. In reality, vaccine hesitancy tends to cluster within specific social groups [9], underscoring the need to understand how unvaccinated children interact with each other [10]. Apart from hindering efforts to eliminate measles, these outbreaks pose immediate risks to vulnerable populations, including young children who can’t yet receive vaccinations and people with certain medical conditions.

This phenomenon of clustering in vaccine hesitancy within social groups is particularly clear in the Netherlands, where members of the Orthodox Protestant Church and the Anthroposophic community tend to have lower MMR vaccine uptake [11,12]. This clustering of susceptible children has given-way to a number of measles outbreaks since introduction of MMR vaccine in 1989 with the largest occurring in 1999 and 2013, each of these with an estimated total infection count of c. 30,000 with large portions of the country affected [13], in both outbreaks the majority of infections reported were in children, with 77% of cases in 2013/14 amongst 4 to 17 year olds [14].

In the Netherlands, over two-thirds of schools are associated with one of 27 denominations, most of which have religious or philosophical affiliations. An analysis of vaccine coverage in Dutch schools in 2013 showed that schools linked to two specific denominations – Anthroposophic and Orthodox Protestant – had a higher concentration of unvaccinated children who had received at least one dose of the MMR vaccine [15]. This finding aligns with recent outbreaks, however, variation in outbreak sizes [16–20] suggest that factors beyond clustering of unvaccinated children within schools also influence outbreak risks. Concretely, clustering of unvaccinated children within individual schools can explain the smaller locally concentrated outbreaks like the outbreak in Anthroposophic schools in 2008 [17], but is not sufficient to explain the very large and geographically disparate outbreaks witnessed in 1999 and 2013 [14,21].

We suggest that focusing solely on vaccine uptake within schools overlooks the potential impact of interactions among unvaccinated siblings. Families may choose schools based on shared affiliations, leading to larger clusters of susceptible individuals that cover broader geographical areas than just single school boundaries. To understand this clustering better and assess its implications for larger outbreaks, we propose a method that examines connections between schools and households using national school registration data. This approach builds on previous research showing that households and schools are primary locations for close and lasting contacts, which play a significant role in disease transmission [22,23]. We have previously shown the effectiveness of using school-household data to study how the structure of the education system relates to the spread of infectious diseases among school-age children [24]. This approach’s reliability has also been confirmed through analysing data related to COVID-19 in the Netherlands [25].

Similarly to Munday et. al. 2021 [24] we construct a comprehensive network that links schools and households with unvaccinated individuals. For this we have used data from the Dutch Ministry of Education to establish household connections between schools. We first analysed the network to establish the connectedness of schools with the same affiliations by quantifying homophily indices, comparing networks with geographical distances between schools and by evaluating the nature of clusters identified using community detection. We then extended our approach to include vaccination informed by vaccine uptake estimates at the school level from Klinkenberg et. al. [15], we can better understand how unvaccinated children interact. To validate the usefulness of this network for assessing outbreak risks and their potential impact, we simulated measles outbreaks within this network and compared the results to a real outbreak that occurred in the Netherlands in 2013/14 [13]. To assess the robustness of this combined network we compare our results with two alternative combined network structures, the first in which MMR uptake is not clustered by school but by postcode, and the second schools are not clustered based on school-households contacts but based on geographical distance.

## Methods

### School network

To construct a network of schools as connected through household contacts (Figure 1A) using the approach we described in Munday et al. 2021 [24], we used government data to calculate the total number of unique contact pairs between schools for the reference date of 1^st^ October 2013 (the year of the last major measles outbreak in the Netherlands). The Ministry of Education (Dienst Uitvoering Onderwijs, DUO) holds data on school attendance for each child in non-private education for the Netherlands (>99% of school aged children). From these data, the ministry calculated the number of children per individual school at each unique address (irrespective of class, age and gender). For each unique address, based on the number of children per school, the number of unique contact pairs between each pair of schools represented in that address was calculated, where a contact pair is a pair of students who live in the same address but attend different schools (for example 2 children school A and 2 children school B form 2×2=4 unique contact pairs between school A and B) (Figure 1B). Subsequently, for each pair of schools in the data, we calculated the total number of contacts across all addresses, resulting in a total number of unique contact pairs between each pair of schools (for example 46 contact pairs between school A and B). In this calculation, schools are defined as school-locations identified by their location number in the DUO database. Therefore, we distinguish between the multiple (geographical) school buildings/locations even though they belong to the same school administration.

**Figure 1.**
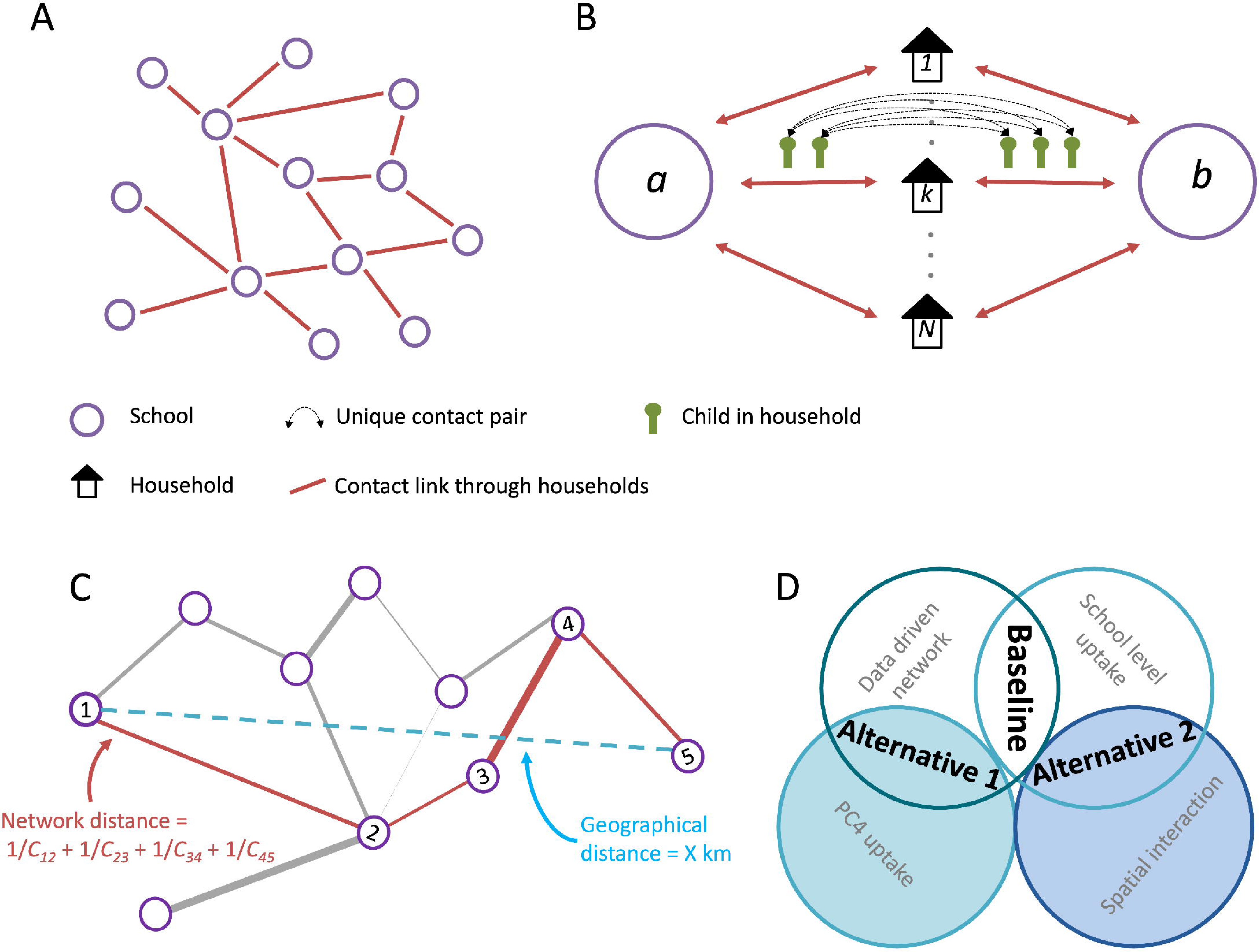
A) Network of schools constructed such that schools are connected when contact is made between pupils of different schools within a household. B) The strength of contact between schools is quantified by calculating the number of unique contact pairs (one child in each school). The number of pairs per household is the product of the number of children who attend school *a* and the number of children who attend school *b*. The total number of unique pairs is the sum of unique pairs in each household with children attending both school *a* and *b*. Figure adapted from [24]. C) Calculation of network distance between nodes 1 and 5 is the sum of the edges along the shortest path between those nodes. D) Schematic of the components of the different network models. The baseline used the data driven contact network and school level uptake. Alternative models: 1. Uptake based on four digit postcode areas of the children who attend the school. 2. Interaction based on a spatial interaction kernel.

### School data

The denomination (see table in supplementary material), the total number of students per school (also on October 2013) and the catchment of each school, the number of students per 4-digit postcode (PC4) was obtained from open source education data [26]—Dutch postcodes are an alphanumeric code of four digits followed by two letters. The exact geographical location of each school was obtained from the full address and postcode of the school site.

### Community structure in the school network

Firstly, to evaluate the connectedness of schools irrespective of denomination, we evaluated the community structure within the school network. Communities represent groups of schools that are more connected to each other than to other schools on the network. For this we used the modularity maximisation based Leiden algorithm, implemented in the *leidenalg* python package [27]. This approach partitions the network such that groups of schools are identified, which are more connected to each other than schools that belong to other groups. The size of the groups detected by the leiden algorithm can be adjusted using the resolution parameter—the higher the value of resolution parameter, the smaller the communities detected (Supplementary material). We first evaluated partitions made using different values of the resolution parameter. To establish the most meaningful scale of communities, we partitioned the network with values of resolution between 0.1 and 1. We evaluated the partitions against four metrics: *Internal edge density[28]*, *Modularity density [29]*, *Neman Girvan modularity [30]* and *Surprise[31]*, for details see supplementary material. We established that a resolution parameter of 1 gave the best score in all metrics and therefore proceeded with this for the further analysis. To establish a consensus partition we generated 20 partitions of the network. From the partitions we calculated a similarity matrix, where each element was equal the the frequency with which each pair of schools was partitioned into the same community. We repeated the process again but instead partitioned the network described by the similarity matrix. We repeated this process until all 20 partitions were identical, we present this partition as the consensus partition. We evaluated each stage of the process by calculating the normalised mutual information of each pair of partitions. We then evaluated the composition of each of the communities in the consensus partition.

To evaluate patterns by geographical location and affiliated denomination in the initially generated partitions, we calculated the mean pairwise probability that any two schools of the same particular denomination or province fall into the same community over the partitioned networks, giving the propensity for schools of particular denominations to form communities in the network.

### Network analysis

We explicitly evaluated the relative connectedness of schools with other schools of the same denomination on the network by calculating the Basic Homophily (*H_i_*) and Coleman Homophily Index (*IH_i_*) [32]; equations 1 and 2) of each school denomination This measure gives the proportion of neighbouring schools that are of the same identity relative to the prevalence of the identity in the school system.

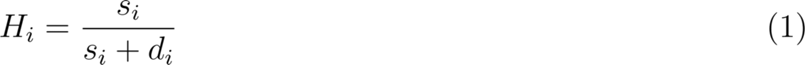

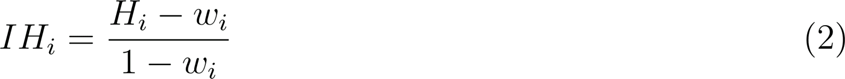

Where, *s_i_* is the number of connections between schools in denomination i, *d_i_* is the total number of connections between schools in denomination i and any other school in the network. and *w_i_* is the proportion of schools in the network that belong to denomination i.

To explore longer-range connections in the network, we compared geographical and network distance between pairs of schools in the network, similarly to previous work by Donker et al[33] studying hospital networks. Network distance was defined as the length of the shortest path between schools on the reciprocal contact network. The weights of the edges in the reciprocal network are equal to the reciprocal of the number of unique contact pairs between schools. The network distance between two schools is therefore the lowest possible sum of edges that form a path between the schools on the reciprocal network (Figure 1 C). I.e. 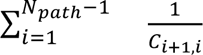 where *C*_*i*+1,*i*_ is the number of contact pairs between consecutive schools I and i+1 in a shortest path of *N_path_* edges.

We calculated the network distance (distance ratio) and geographic distance (km) of 1000 randomly sampled pairs of schools from the biggest faith-based school denominations in the Netherlands: Roman Catholic (Rooms-katholic) and mainstream Protestant (Protestants-Christelijk). We also calculated the distances for Orthodox-Protestant and Anthroposophic (Antroposofisch) faith identities, which are most closely associated with low vaccination uptake.

Schools with a low distance ratio are more closely connected on the network relative to their geographic distance than schools with higher distance ratios. We calculated distance ratios for pairs of schools of the same faith identity to that of schools in the rest of the network, defined as all schools not associated with that faith identity. To account for geographic location of schools, we compared distance ratios for schools sampled from the ‘rest of the network’ from the same two-digit postcode area as each school sampled from the denomination of interest.

### Transmission model

In addition to our analysis of the network of schools, we evaluated the epidemiological relevance of any increased connectedness between schools, specifically concerning outbreaks of measles. We used the network to simulate outbreaks of measles in school-aged children (supplementary information). We extended the method used in Munday et al. [24] to include vaccination. In a generation-based model, each school could be in one of three epidemiological states: Susceptible, Infected or recovered. Schools in the susceptible state had immunity amongst its pupils equal to the complement of the estimated vaccination coverage (*V_j_*) in that school in October 2013 calculated by Klinkenberg et al [15] (Figure S3). When a school becomes infected, the total outbreak size within this school is obtained by a final size equation [34] (equation 3) considering the total number of students, the number of susceptibles and *R_0_*.

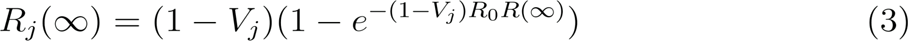

Subsequently an infected school, *j*, can infect connected school, *i*, with probability *P_trans,i,j_* depending on the number of contact pairs (*C_ij_*), the probability that a contact from the infected is infected (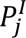) (based on the final outbreak size), the percentage susceptible in the connected school, 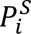, and the probability that introducing an infected child into the susceptible school would lead to an outbreak (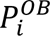) (see supplementary material for full description).

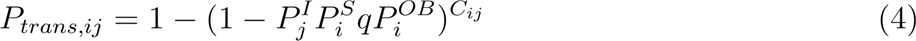

For our application, we have assumed a gaussian offspring distribution for within-school transmission, hence 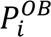 is equal to 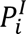. Outbreaks were simulated from an initial state, where one or more schools were in the infected state and the remaining schools were in the susceptible state. We set the “within school” reproduction number (*R_0_*) to 15, consistent with estimates for measles (12-18) [35] and the probability of transmission to a susceptible sibling (*q*) of 0.5.

### Alternative network models

To evaluate the importance of the specific connectedness in the school network and vaccination at school level to the simulated outbreaks, we designed alternative models which contained only part of the information used in the full model (Figure 1 D).

“Alternative model 1” was designed to establish the importance of clustering of unvaccinated children in particular schools. We analysed an alternative parameterization assuming children had a probability of being vaccinated equal to the vaccine uptake of the PC4 where they lived. We used data on the residence of children in each school to calculate the proportion of children in each school who live in each PC4. School vaccination uptake was set as the weighted average of vaccination uptake at PC4 level, weighted by the proportion of children who live in each PC4.

We included “alternative model 2” to assess the importance of the specific network structure defined by the school data to the overall dynamics of outbreaks. We constructed an alternative school contact network where the geographic distance between connected schools followed a similar relationship to the baseline model, however contact is spread evenly over all schools according to that relationship. The spatial distribution of a school’s immediate neighbours in the data-based school network can be described by an exponential distribution. We therefore used this relationship and weighted by the degree of the connecting schools to describe the spatial relationship for alternative model 2. To calibrate the spatial model, we matched the distribution of distance between schools connected by contact pairs to the school data derived network (Details in the supplementary material).

### Outbreak simulations

In our model each school can occupy one of three states: susceptible, infected, or recovered. Susceptible schools are represented as initially described on the transmission probability network, with immunity profile equal to 1 – *V*. Infected schools are those affected by an outbreak, and have a probability of infecting neighbouring susceptible schools (*P_trans,i,j_*) as defined above. After an outbreak occurred, we assumed that the school had effectively depleted its susceptible population, entering the recovered state where the school could not be re-infected. For each iteration of the model we sampled a set of vaccine uptake values (per school) from the distributions given by Klinkenberg et. al (2022). For each sample the values of *P_trans,i,j_* collectively form a static directed network of transmission probabilities. To create outbreak realisations, we sampled edges of an equivalent network where edges have a weight of 1 with probability *P_trans,i,j_*. These binary directed networks represent paths along which transmission can occur in the simulation. For each realisation of an outbreak, one or more schools were set to be in the infected state, outbreaks were then described by trees described by the successive out-edges of the binary network.

### Evaluating the model against epidemiological data

To evaluate the ability of the network-based simulations to capture observed measles epidemiology, we compared simulated outbreaks to final estimates of cases from a large outbreak in 2013/14. To do so, the cumulative measles cases per PC4 for the 2013 - 2014 were obtained from the National registry of reportable infectious disease (OSIRIS).

Simulations of the outbreak were initiated by placing the two schools, which are believed to be the index-schools in the outbreak of 2013/14, in the infected state. All other schools were initiated in the susceptible state. We calculated the total number of students expected to be infected per school by multiplying the final size proportion estimate by the number of students in the school. Then we allocated infected students proportionally to PC4-areas, based on the proportion of children in each school that live in each PC4 area, which allowed us to compare the simulated outbreaks to the observed historical outbreak. We ran the model 10,000 times and calculated the mean number of cases per PC4 area over all realisations of the simulation. Average model outcomes were compared using Receiver Operating Characteristic (ROC) on PC4-level, based on the presence or absence of cases. To reflect the relative importance of PC4s with higher reported or simulated incidence, we also calculated a weighted ROC (wROC). For this measure, when calculating sensitivity, each PC4 with cases reported is weighted by the proportion of all cases reported in that PC4. Hence, whereas for the unweighted ROC the sensitivity value is the proportion of areas with cases reported that also had cases predicted, for the wROC the sensitivity value is the proportion of cases reported that occurred within PC4s where cases were predicted by the model.

### Evaluating the risk of outbreak posed by each school

We used the transmission model described above to evaluate the relative risk an outbreak originating in each school poses to the network as a whole. To quantify this risk, we simulated (as described above) 1000 outbreaks initiated at each school in the network and reported the mean number of schools and children infected.

To establish the relevance of the specific schools independent of their vaccine status we present the number of schools and children infected by a school by that school’s vaccine uptake. To evaluate the sensitivity of these results to the network structure, we compared results from our transmission model with those of Alternative Model 2 (with a spatially-derived network).

All analysis was performed in Python 3 [36]. Network analysis was performed using NetworkX [37], community detection was performed using the Leidenalg [27] package and evaluation of the partitions was performed using the CDLIB [38] package. All other analyses made use of the scientific python package library [39].

## Results

### Communities in the network

We found that the algorithm converged on a consensus partition after two rounds (Figure 2), this attests to the stability of the initial set of partitions. Indeed, after the first round only 2 unique partitions were found, which themselves were very similar, with a normalised mutual information score of 0.98.

**Figure 2.**
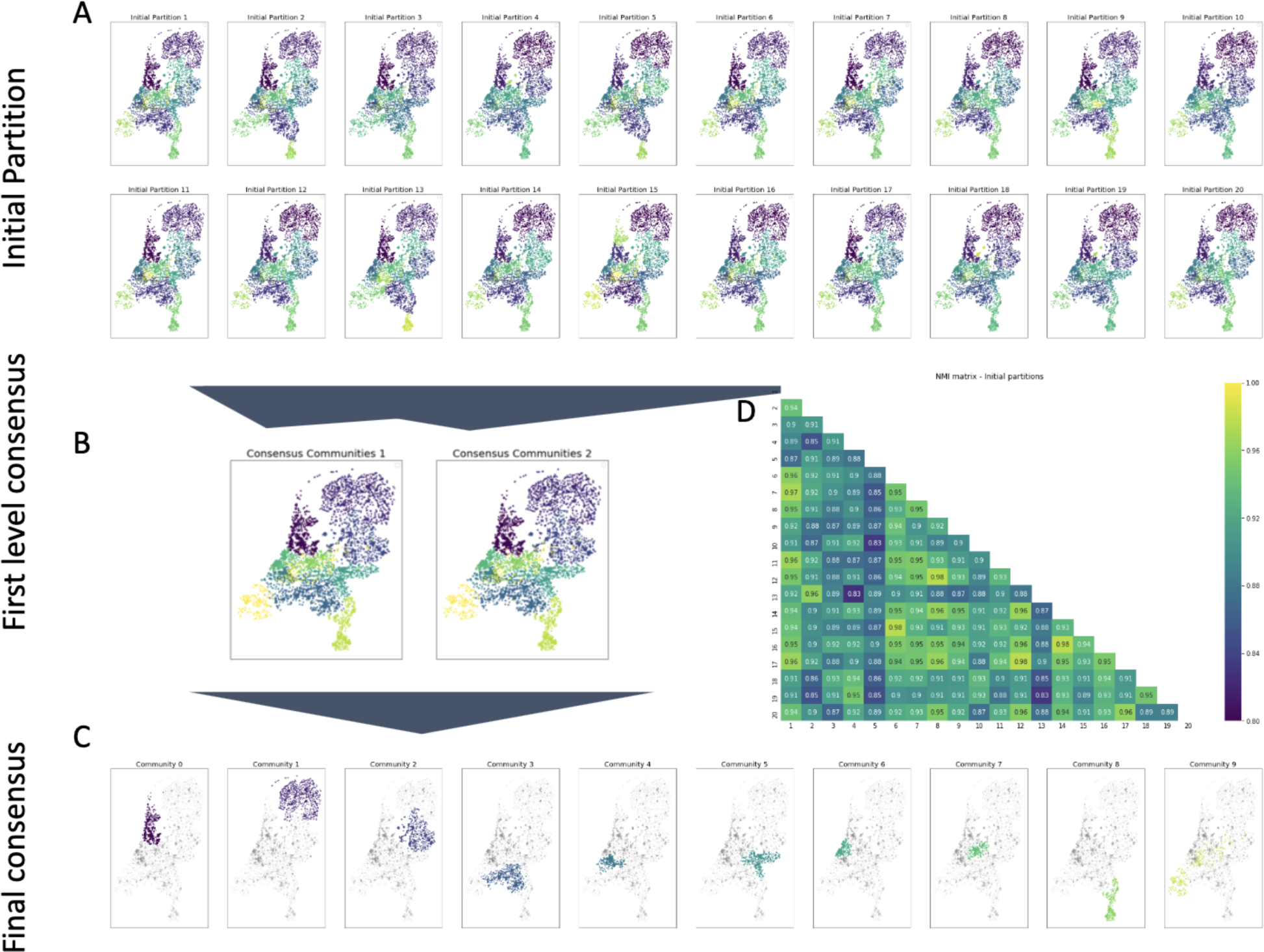
Consensus partition - Community structure of the school network. A to C show the locations of schools in the Netherlands, the colour of the markers indicates the community of schools in the partition. The panels in A show the 20 initial partitions, B shows the partitions from the first round of the ensembling algorithm. C shows each community in a separate panel in the final consensus partition, grey points show the locations of other schools (not in the community in that panel). D shows a matrix normalised mutual information (NMI) between the initial partitions.

The final consensus partition resulted in mostly geographically organised communities, with high probability of schools in the same province being assigned the same community. In general, any preference of connection between schools of the same religious affiliation was not sufficient to overpower the strong geographical component in the communities. The exception was community 9 (Figure 3), which was partially associated with the province of Zeeland (250 schools, 61% of the community). However, an additional 155 schools from other provinces were included in this community including 129 (31% of the community) from the protestant orthodox (Reformatorisch) denomination. In total 166 (41% of of the community) schools associated with the protestant orthodox denomination were included in this community from 6 different provinces, this represents 79% of all schools of this denomination.

**Figure 3.**
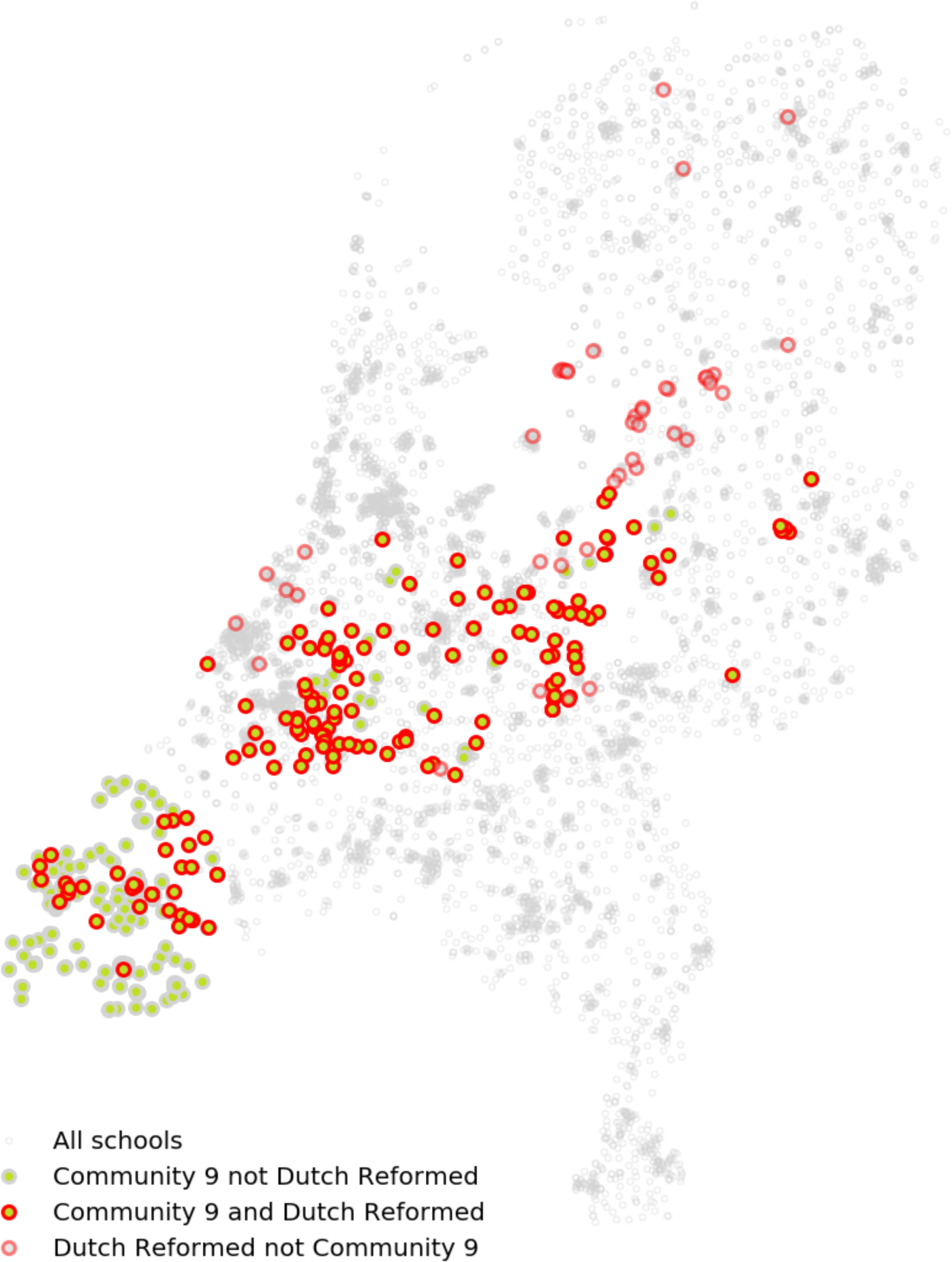
Consensus partition community 9 which is primarily composed of schools in the Zeeland province and Protestant Orthodox schools (Reformatorisch) spread across the Nation.

The pairwise probability that schools of the same province fell into the same partitioned communities was high with a mean of 0.75 (Table 1). In contrast the mean pairwise probability that schools of the same denomination were partitioned into the same communities was much lower with a mean of 0.28. There were two denominations which were excluded from the analysis, Jewish schools (Joods) and Moravian Church (Evangelische broedergemeenschap) schools, which both have only a small number of geographically clustered schools in the network resulting in pairwise probabilities of 1.0. Of the remaining, larger denominations, the Protestant Orthodox (Reformatorisch) schools had the highest pairwise probability of 0.55. In contrast, Athroposophic schools (Antroposofisch), the other denomination associated with low MMR uptake, had much lower pairwise probability of 0.12. (Table 1)

**Table 1.**
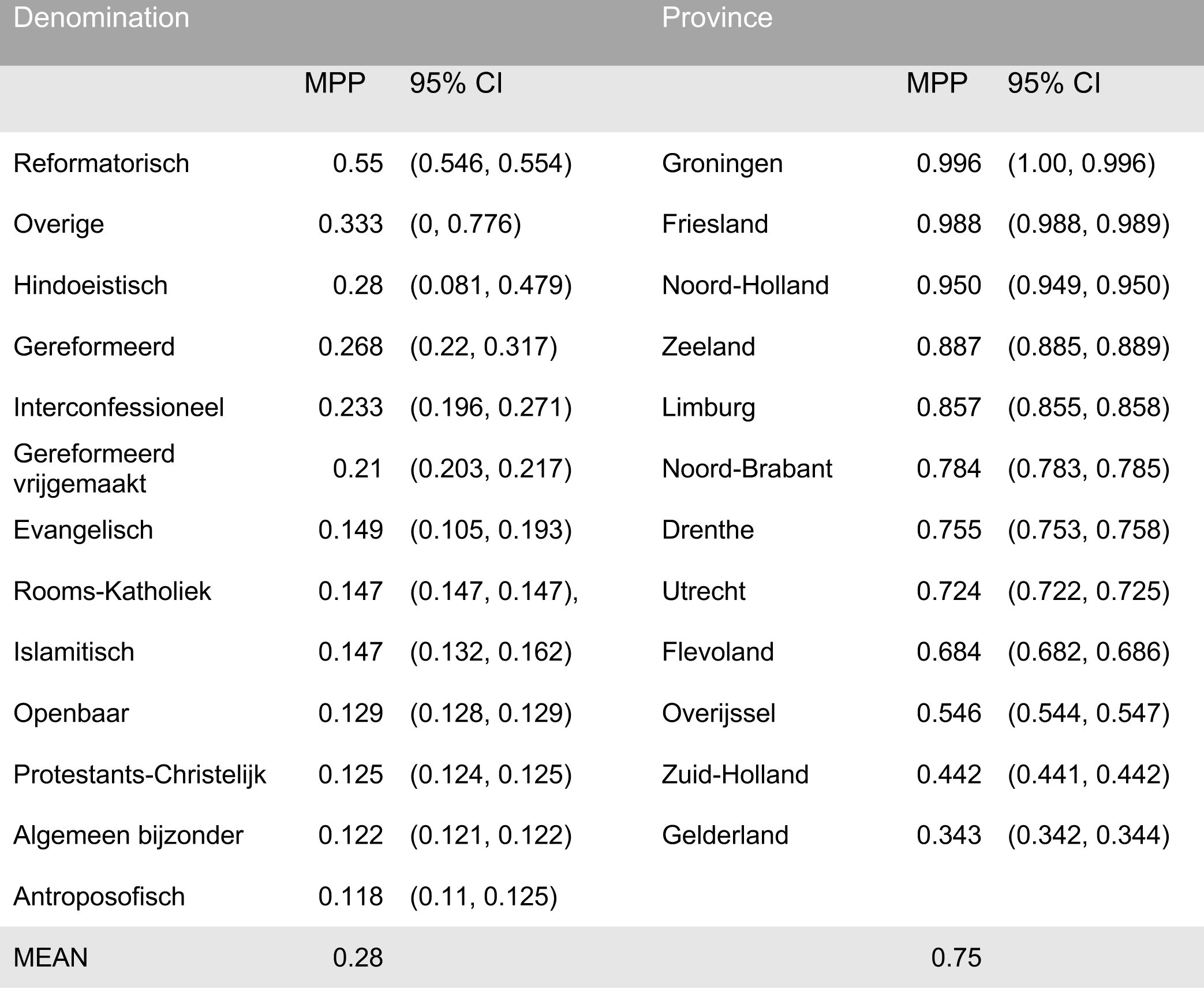
Mean pairwise probability of being partitioned into the same community by School denomination and Region.

### Network analysis

On average each school was directly connected to 39.8 other schools through 225.9 contact pairs. These values were much lower for primary schools, 27.5 and 122.1 respectively. Conversely secondary schools had many more immediate neighbours with 109.7 connected schools through 813.9 contact pairs (figure 4 A). Orthodox Protestant (Reformateorisch) schools tended to have a higher ratio of contact pairs to the number of schools compared to the rest of the network, whereas Anthroposophic (Antroposofisch) schools had a more characteristic relationship between the number of neighbouring schools and contact pairs indicating more thinly distributed contact between a larger number of schools.

**Figure 4.**
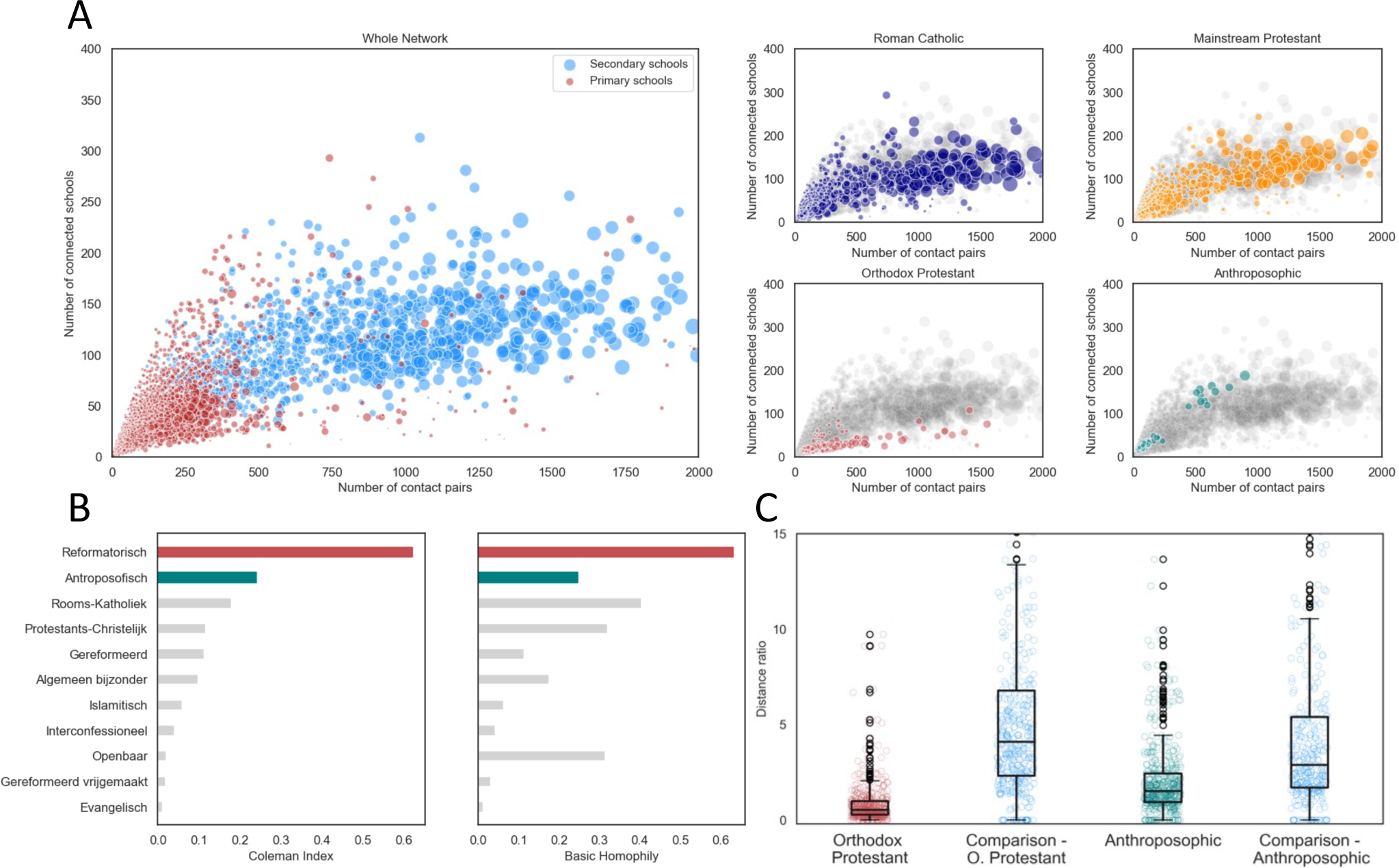
A) The number of contact pairs connected to other schools against the number of connected schools for each school in the network. The main panel shows the primary schools in red and secondary schools in blue, the four panels on the right show the two largest school denominations (protestant and catholic) alongside the Orthodox Protestant and Anthroposophic denominations, the rest of the schools in the network are shown in grey for comparison. B) The 11 faith identities with the highest Coleman Homophily Index (CHI). in the left panel bars show the CHI of each faith identity. In the right panel, bars show the basic homophily of each faith identity. Red bars highlight the Orthodox Protestant (Reformatorisch) and Anthroposophic (Antroposofisch) identity schools, where vaccination uptake is known to be low. C) Boxplot of distance ratio for pairs of Orthodox Protestant (Reformatorisch) [Here labelled “Reformed”] and Anthroposophic (Antroposofisch) schools and geographically equivalent sample from the rest of the network.

### Relative connectedness

For the majority of faith identities there is a positive homophily index, suggesting that households are more likely than expected to have children in two or more schools of the same faith identity than would be expected at random (figure 4 B).

The four faith identities with the highest CHI were Orthodox Protestant, Athroposophic, Roman Catholic and Mainstream Protestant, With CHI ranging from 0.62 to 0.12. Notably the two school identities with the highest Coleman Index were Orthodox Protestant (Reformatorisch) (Reformatorisch) (BH = 0.63, CHI= 0.62) and Anthroposophic (Antroposofisch) (BH = 0.25, CHI = 0.24), which are the two faith identities with populations who systematically refuse vaccination (Figure 4).

### Distances across the network

The mean ratio of network to geographic distance was 3.09 pairs^−1^ km^−1^ for the whole network, 0.54 pairs^−1^ km^−1^ for Orthodox Protestant (Reformatorisch), 3.82 pairs^−1^ km^−1^ for Anthroposophic (Antroposofisch), 3.75 pairs^−1^ km ^−1^ for Roman Catholic and 3.17 pairs^−1^ km^−1^ for mainstream Protestant. This indicates that The Orthodox Protestant (Reformatorisch) faith identity alone forms extended chains of schools strongly linked through households, whereas the other faith identities are generally as connected as any schools in the whole network.

The scatter plots of network distance against geographic distance revealed that in both cases network paths were shorter between the Orthodox protestant (Reformatorisch) and Anthroposophic (Antroposofisch) schools than between randomly selected geographically equivalent schools.

The distance ratio (network distance divided by geographic distance) distribution was lower for Orthodox Protestant (Reformatorisch) schools and Anthroposophic (Antroposofisch) schools than their comparison samples. With mean distance ratios of 0.54 pairs^−1^ km ^−1^ and 3.83 pairs^−1^ km ^−1^ for Orthodox Protestant (Reformatorisch) and Anthroposophic (Antroposofisch) schools respectively and 5.18 x 10^−3^ and 4.81 x 10^−3^ for their respective comparators (Figure 4).

### Simulation studies

Using the baseline model (National school data contact network and school level vaccine uptake estimates), 1000 outbreak simulations with the initial schools set to the two schools first identified in the 2013 outbreak resulted in a mean of 23,497 (23,340 – 24,267 IQR) infections. The geographical distribution of cases was broadly consistent with the reported cases in 2013/14. There was a high likelihood of cases being reported in PC4 areas in the centre of the country and the southwest. There was also a high likelihood of infections in a small region in the north of the country (Figure 5).

**Figure 5.**
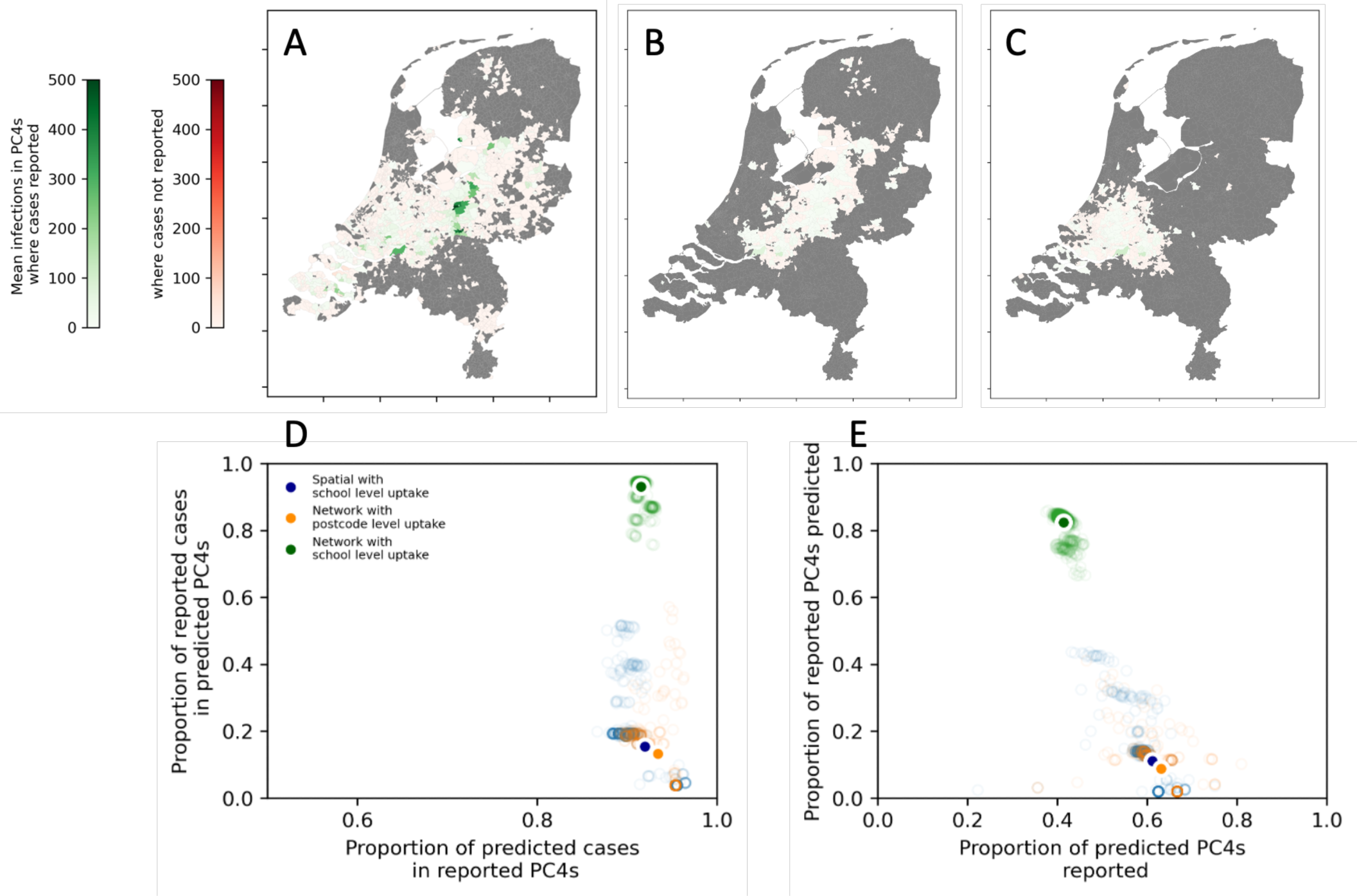
Mean number of cases across 1000 simulated in each PC4 region with a reporting rate of 10% (from estimates in literature). A) the baseline model: School data network with school level uptake., B) Alternative model 1: School data network with PC4 level uptake, C) Alternative model 2: Spatial network with school level uptake D) weighted sensitivity and specificity and E) unweighted sensitivity and specificity of the baseline and alternative network models.

When Alternative model 1 was used, (national school data in combination with the vaccination uptake in schools estimated from PC4 level vaccine uptake), the mean final size of the outbreaks was 259 (25 – 313 IQR). The cases were distributed in a narrow strip, with a high frequency of cases in the region from the south west region to the north east of the central region (Figure 5).

When Alternative model 2 was used (the spatially derived contact network and school level vaccine uptake estimates), the final size of the outbreak was 1241 (207 – 1619 IQR) cases. The majority of cases predicted occurred in schools in the central region of the country, with low probability of detecting infection in any other regions (figure 5D).

Using the unweighted ROC, the mean sensitivity (proportion of PC4s where cases reported that were predicted by the model) was 0.84, 0.28 and 0.18, for the baseline model, alternative model 1 and alternative model 2 respectively (Figure 5D). The mean specificity (proportion of PC4s where cases were predicted that also had cases reported) was 0.40, 0.54 and 0.57 for baseline model, alternative model 1 and alternative model 2 respectively.

Considering the weighted ROC. The mean sensitivity was 0.94, 0.38 and 0.23 for the baseline model, alternative model 1 and alternative model 2 respectively (Figure 5E). The mean specificity was 0.94, 0.91 and 0.91 for School data network with school vaccination, Spatial network with school vaccination and School data with PC4 level vaccination respectively.

### Outbreak size by school where the outbreak is initiated

The overall risk posed by an outbreak in each particular school was quantified by finding the distribution of final outbreak size. For both the school data and spatial networks, the majority of schools had a very low mean outbreak size as no sustainable transmission was observed in any simulation.

For the full network model, the maximum mean outbreak size was 145 schools and 23,505 children and was associated with a Orthodox Protestant (Reformatorisch) school (Figure 6). In general, Orthodox Protestant (Reformatorisch) schools generated large outbreaks and were particularly high for schools with very low vaccination coverage. Outbreaks seeded in Anthroposophic (Antroposofisch) schools generally remained much smaller, with a maximum of 3 schools and 462 children.

**Figure 6.**
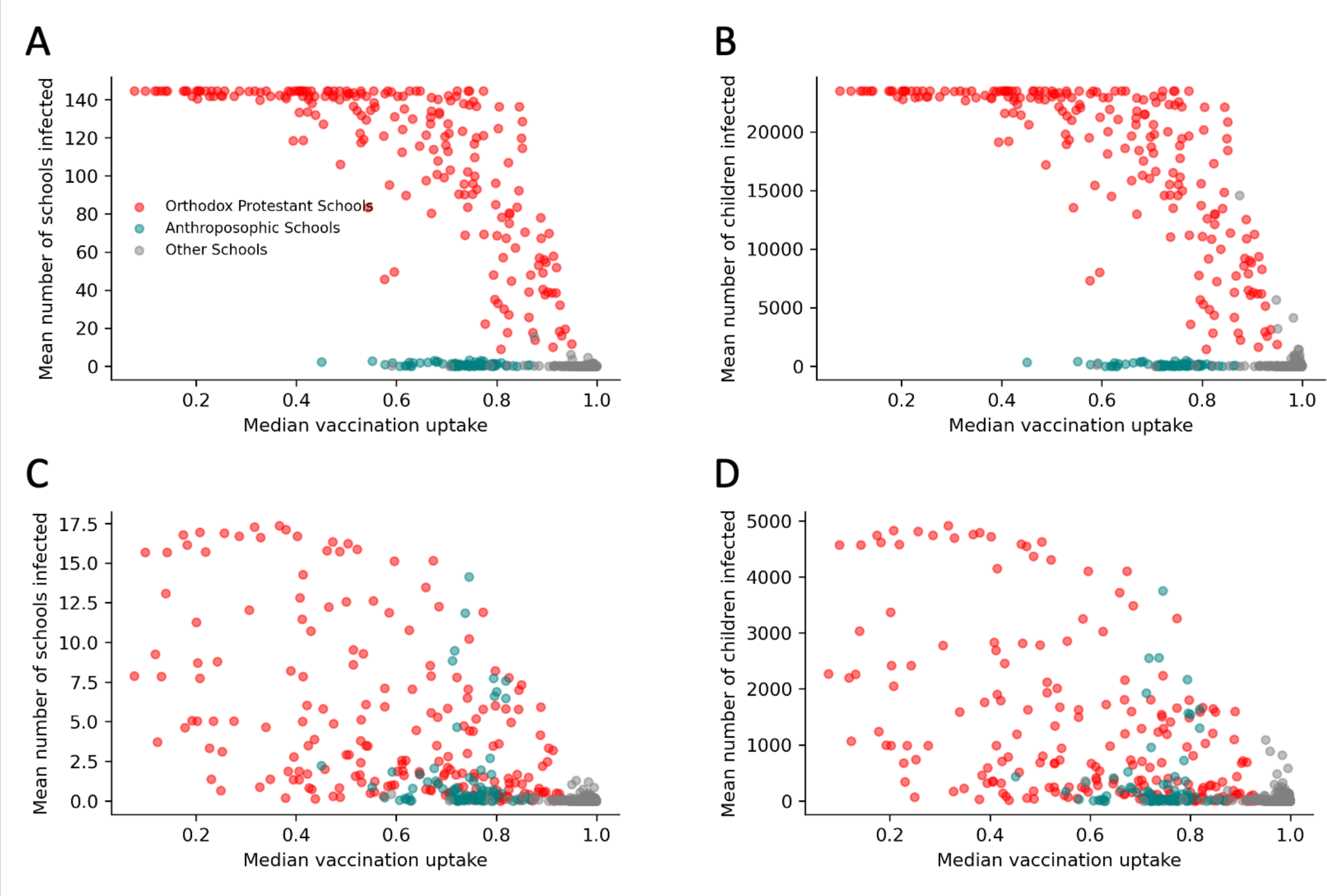
Mean outbreak final size by school where outbreak is seeded. Red points indicate Orthodox Protestant (Reformatorisch) schools, green points indicate Anthroposophic (Antroposofisch) (Antroposofisch) schools, grey points indicate other schools. A) School data derived network-mean number of schools infected, B) School data derived network-mean number of children infected, C) Spatially derived model-mean number of schools infected, D) Spatially derived model-mean number of children infected.

Outbreaks simulated on alternative model 2, with a spatially derived network, were generally much smaller. There was a trend with vaccine uptake, however there remained schools where vaccine uptake was low that still only seed very small outbreaks. The schools that seeded the largest outbreaks using alternative model 2 had a mean of 17 infected schools and 4,916 children infected. The Orthodox Protestant (Reformatorisch) schools seeded the largest outbreaks. However, the difference between Orthodox Protestant (Reformatorisch) and Anthroposophic (Antroposofisch) schools was much less substantial than for the full network model with a number of Anthroposophic (Antroposofisch) schools seeding outbreaks greater than that predicted by the model derived from the school data, with a maximum of 14 schools and 3,751 children. Notably the some Anthroposophic (Antroposofisch) schools seeded outbreaks comparable to those seeded by Orthodox Protestant (Reformatorisch) schools with similar vaccine uptake.

## Discussion

We used a national school-household network including primary and secondary schools that represent >99% of school aged children in the Netherlands as a framework to quantify the outbreak risk of measles given observed vaccine-uptake by school. Doing so revealed that large close networks exist for specific faith identities, where the Orthodox Protestant schools connect households (and vice versa) over a large geographical distance in the Netherlands. Our network approach (parameterized on 2013 data) was able to predict accurately (sensitivity and specificity of both 0.94) the measles cases per PC4 level as observed in the 2013/14 outbreak in the Netherlands. The total incidence predicted by the model (approx. 23,500 infections) was markedly close to the estimates from literature (30,000 infections with 77% in school-aged children (4-17 years old)). This demonstrates that by incorporating a school-household network on top of school-specific uptake data we can quantify outbreak-risks for measles, the overall outbreak size, the geographical spread of these outbreaks, and identify which particular school contributes to which degree to observed cases per PC4. Our analysis can be repeated on an annual basis utilising the data held in the vaccination registry, and administrative data of the department of education, so as to assist national and local public health teams in their assessment of risk and subsequently which groups of parents/children to target for national and local campaigns to reduce this risk.

Our analysis of the network of schools reveals that Orthodox Protestant are much more connected via households than would be expected geographically, evidenced by short network distances when compared to geographical distances. This property is not shared with Anthroposophic schools. The increased connectedness of Orthodox Protestant schools combined with low coverage of MMR creates large clusters of unvaccinated children, which would be expected to increase the risk of large outbreaks of Measles, Mumps and Rubella amongst this population. Our simulation studies allowed us to quantify the implications of the clustering we identified in the network analysis. Our results have important implications for understanding the determinants of large outbreaks of measles in the Netherlands and their timing.

Firstly, our findings indicate that the distribution of unvaccinated children within particular schools, and the specific links between these schools greatly increase the potential for large outbreaks to occur. With these factors accounted for in the model, outbreaks similar to that observed in 2013/14 can be simulated by accounting for school and household transmission only. This finding suggests that, in a population with immunity provided only by vaccination, outbreaks have a clear determinable reach, which is not reliant on chance encounters or rare long-range transmission events.

Secondly, our evaluation of the school-network highlights the difference between orthodox protestant and anthroposophic schools in how they interact with schools of their own denomination compared to the rest of the network. This provides a mechanistic explanation for the difference in outbreaks observed in the orthodox-protestant and anthroposophic communities in the past, where those in anthroposophic communities have remained relatively small (a small number of schools), whereas the outbreak in 2013 (largely in the orthodox-protestant community) was much larger [17,20].

Thirdly, it is evident from the degree distributions of primary and secondary schools that secondary schools are more connected on the network, this is consistent with observations of the school network in England, UK. This suggests that secondary schools may play a more substantial role in determining the spatial distribution of measles outbreaks than primary schools - a property that could be explored further in future work.

Further, since the variation in outbreak size is due to structural differences in the population, it is likely that future outbreaks in these communities would follow similar patterns, if the structure of the school system remains comparable in years to come.

Our approach made some important simplifying assumptions that the majority of transmission of measles between children occurs between contacts that either reside in the same home or attend the same school.

First, the model made some assumptions about transmission routes in the population. The model does not account for possible transmission between children outside school and household (e.g. sport, church activities, or assumes that these contacts can still follow school networks) and the model does not take into account transmission outside of the school-aged population. Adults and preschool-age infants are likely to contribute to transmission to some degree. These neglected routes of transmission could potentially influence transmission dynamics in a way that this model cannot capture. In 2013/14 there were 438 cases (19%) in children between 1 and 4, lower than the 819 (30%) and 868 (32%) cases in 5-9 and 10-14-year-old age groups respectively, suggesting less transmission within pre-school age than school aged children[20]. The presence of pre-school institutions in the network would provide additional connectivity on the network which may increase transmission opportunities, particularly between primary schools, however since the contribution of primary schools to connectivity in the network, it might be expected that pre-school settings, which tend to be smaller would provide limited additional transmission opportunity compared to the current network.

Secondly, our model does not simulate within-school transmission dynamics, but instead assumes a deterministic final size approximation [34], which occurs with a probability determined by the effective reproduction number in that school. This cannot capture the contribution of outbreaks that do not reach sustainable transmission within schools, but still represent some small risk in terms of infecting other schools with the few pupils that are infected.

Finally, the model works purely on a ‘generational’ basis, with no explicit temporal element. This restricts its use to modelling the overall incidence of an outbreak without modelling the temporal dynamics. It is the case that transmission would only occur through other routes (not schools) during school holidays and weekends, for example. This cannot be captured in simulations with the model in its current form.

These limitations, however, do not detract from the findings that the school system provides a system of contact that can facilitate large outbreaks amongst unvaccinated children in the Orthodox Protestant (Reformatorisch) population but not amongst children in Anthroposophic schools.

Further analysis of this network could allow study of other infectious diseases such as mumps and rubella, which are also prevalent amongst school-aged children within the same socio-religious populations. The model could also be extended to analyse outbreaks of influenza, where a large degree of transmission occurs within school age children. Another use of this framework could be to evaluate the effectiveness of various other intervention strategies, such as school closure. This method could also be applied in other settings where vaccine uptake is strongly related to particular social groups [9], this relies on detailed school data being made available.

## Conclusion

Our results indicate that important correlations between religious faith and vaccination refusal in the Netherlands may lead to large clusters of children who are at high risk of measles infection, through school and household contact. We found that groups associated with low vaccine uptake, the Orthodox Protestant Church and the Anthroposophic community displayed substantial homophily on the school network, indicating higher degree of connectedness than to other schools. By explicitly modelling connections, we can provide important insights into the epidemiology of measles in the Netherlands and why it may vary between socio-religious groups. The results of our simulation studies suggest that the network improves our model’s ability to describe observed epidemiology from previous outbreaks. High and long-range network connectedness between Orthodox Protestant schools revealed in our network analysis leads to larger outbreaks in this community compared to Anthroposophic schools, between whom connectivity on the network is weaker. This framework provides a basis for evaluating risk of large outbreaks of measles in the Netherlands and could be further developed to aid future outbreak response.

## List of abbreviations

DUO: the Education Executive Agency in the netherlands
HI / CHI: Homophily index / Coleman Homophily Index
IQR: Interquartile Range
MMR: The Measles, Mumps and Rubella Vaccine
MPP: Mean Pairwise Probability
OSIRIS: The Netherlands National registry of reportable infectious disease
PC4: Four digit postcode
RIVM: National Institute for Public Health and the Environment
ROC / wROC: Receiver Operating Characteristic / Weighted Receiver Operating Characteristic

## Declarations

The authors declare no conflicts of interest

## Ethics approval

Ethical approval was obtained from the London School of Hygiene and Tropical Medicine (16028-1), this included the use of the data provided by DUO as well as OSIRIS. Data was prepared within the ministry, and only cumulative links per school were shared with us.

## Consent for publication

Not applicable

## Availability of data and materials

The code and school network data is available at github.com/jdmunday/SchoolsMealesNL. The measles case data is available on request. School population statistics are publicly available from: https://www.duo.nl/open_onderwijsdata/index.jsp. The Measles case data is available on request from RIVM

## Authors contributions

JDM, KEA and AJVH, conceived of and planned the analysis; JDM performed the main analysis with supervision from KEA and AJVH and scientific input from KEA, DK, MM, EF, SH, JW and AJVH. MM and EF provided data for the construction of the school network. JDM wrote the manuscript. All authors edited and reviewed the manuscript. All authors read and approved the final manuscript.

## Data Availability

The code and school network data is available at github.com/jdmunday/SchoolsMealesNL. School population statistics are publically available from: https://www.duo.nl/open_onderwijsdata/index.jsp. The Measles case data is available on request from RIVM

https://www.duo.nl/open_onderwijsdata/index.jsp

## Acknowledgements

The authors wish to thank members of the NIHR HPRU in immunisation and CMMID at LSHTM and the Infectious Disease Modelling Unit at RIVM for feedback. In particular the authors would like to thank Mark Jit, Petra Klepac, Sebastian Funk, Ada Collis Munday, Leon Danon and John Read for their insightful discussions.

## Additional Files

Supplementary Figures

## References

1. Roberts L. Is measles next? Science. 2015;348: 958–61, 963.

2. Baugh V, Figueroa J, Bosanquet J, Kemsley P, Addiman S, Turbitt D. Ongoing measles outbreak in Orthodox Jewish community, London, UK. Emerg Infect Dis. 2013;19: 1707– 1709.

3. Stein-Zamir C, Abramson N, Shoob H, Zentner G. An outbreak of measles in an ultra-orthodox Jewish community in Jerusalem, Israel, 2007--an in-depth report. Euro Surveill. 2008;13: 5–6.

4. Cohen BJ, McCann R, van den Bosch C, White J. Outbreak of measles in an Orthodox Jewish community. Wkly releases (1997–2007). 2000;4. doi:10.2807/esw.04.03.01675-en

5. Lernout T, Kissling E, Hutse V, De Schrijver K, Top G. An outbreak of measles in orthodox Jewish communities in Antwerp, Belgium, 2007-2008: different reasons for accumulation of susceptibles. Euro Surveill. 2009;14. doi:10.2807/ese.14.02.19087-en

6. Firman N, Marszalek M, Gutierrez A, Homer K, Williams C, Harper G, et al. Impact of the COVID-19 pandemic on timeliness and equity of measles, mumps and rubella vaccinations in North East London: a longitudinal study using electronic health records. BMJ Open. 2022;12: e066288.

7. Bedford H, Donovan H. We need to increase MMR uptake urgently. BMJ. 2022;376: o818.

8. Funk S, Knapp JK, Lebo E, Reef SE, Dabbagh AJ, Kretsinger K, et al. Combining serological and contact data to derive target immunity levels for achieving and maintaining measles elimination. bioRxiv. bioRxiv; 2017. doi:10.1101/201574

9. Fournet N, Mollema L, Ruijs WL, Harmsen IA, Keck F, Durand JY, et al. Under-vaccinated groups in Europe and their beliefs, attitudes and reasons for non-vaccination; two systematic reviews. BMC Public Health. 2018;18: 196.

10. Liu F, Enanoria WTA, Zipprich J, Blumberg S, Harriman K, Ackley SF, et al. The role of vaccination coverage, individual behaviors, and the public health response in the control of measles epidemics: an agent-based simulation for California. BMC Public Health. 2015;15: 447.

11. Nic Lochlainn LM, Woudenberg T, van Lier A, Zonnenberg I, Philippi M, de Melker HE, et al. A novel measles outbreak control strategy in the Netherlands in 2013-2014 using a national electronic immunization register: A study of early MMR uptake and its determinants. Vaccine. 2017;35: 5828–5834.

12. Klomp JHE, van Lier A, Ruijs WLM. Vaccination coverage for measles, mumps and rubella in anthroposophical schools in Gelderland, The Netherlands. Eur J Public Health. 2015;25: 501–505.

13. Woudenberg T, Woonink F, Kerkhof J, Cox K, Ruijs WLM, van Binnendijk R, et al. The tip of the iceberg: incompleteness of measles reporting during a large outbreak in The Netherlands in 2013-2014. Epidemiol Infect. 2018;147: e23.

14. Woudenberg T, van Binnendijk RS, Sanders EAM, Wallinga J, de Melker HE, Ruijs WLM, et al. Large measles epidemic in the Netherlands, May 2013 to March 2014: changing epidemiology. Euro Surveill. 2017;22. doi:10.2807/1560-7917.ES.2017.22.3.30443

15. Klinkenberg D, van Hoek AJ, Veldhuijzen I, Hahné S, Wallinga J. Social clustering of unvaccinated children in schools in the Netherlands. Epidemiol Infect. 2022;150: e200.

16. Wallinga J, Heijne JCM, Kretzschmar M. A measles epidemic threshold in a highly vaccinated population. PLoS Med. 2005;2: e316.

17. van Velzen E, de Coster E, van Binnendijk R, Hahné S. Measles outbreak in an anthroposophic community in The Hague, The Netherlands, June-July 2008. Euro Surveill. 2008;13. doi:10.2807/ese.13.31.18945-en

18. Hahne S, te Wierik MJM, Mollema L, van Velzen E, de Coster E, Swaan C, et al. Measles outbreak, the Netherlands, 2008. Emerg Infect Dis. 2010;16: 567–569.

19. van den Hof S, Meffre CM, Conyn-van Spaendonck MA, Woonink F, de Melker HE, van Binnendijk RS. Measles outbreak in a community with very low vaccine coverage, the Netherlands. Emerg Infect Dis. 2001;7: 593–597.

20. Woudenberg T, van Binnendijk RS, Sanders EAM, Wallinga J, de Melker HE, Ruijs WLM, et al. Large measles epidemic in the Netherlands, May 2013 to March 2014: changing epidemiology. Euro Surveill. 2017;22. doi:10.2807/1560-7917.ES.2017.22.3.30443

21. van den Hof S, Meffre CM, Conyn-van Spaendonck MA, Woonink F, de Melker HE, van Binnendijk RS. Measles outbreak in a community with very low vaccine coverage, the Netherlands. Emerg Infect Dis. 2001;7: 593–597.

22. De Cao E, Zagheni E, Manfredi P, Melegaro A. The relative importance of frequency of contacts and duration of exposure for the spread of directly transmitted infections. Biostatistics. 2014;15: 470–483.

23. Melegaro A, Jit M, Gay N, Zagheni E, Edmunds WJ. What types of contacts are important for the spread of infections?: using contact survey data to explore European mixing patterns. Epidemics. 2011;3: 143–151.

24. Munday JD, Sherratt K, Meakin S, Endo A, Pearson CAB, Hellewell J, et al. Implications of the school-household network structure on SARS-CoV-2 transmission under school reopening strategies in England. Nat Commun. 2021;12: 1942.

25. van Iersel SCJL, Backer JA, van Gaalen RD, Andeweg SP, Munday JD, Wallinga J, et al. Empirical evidence of transmission over a school-household network for SARS-CoV-2; exploration of transmission pairs stratified by primary and secondary school. Epidemics. 2023;43: 100675.

26. DUO - Open onderwijsdata. In: duo.nl [Internet]. [cited 24 Jan 2024]. Available: https://www.duo.nl/open_onderwijsdata/index.jsp

27. Traag VA, Waltman L, van Eck NJ. From Louvain to Leiden: guaranteeing well-connected communities. Sci Rep. 2019;9: 5233.

28. Ronhovde RKDDRRP, Nussinov Z. An edge density definition of overlapping and weighted graph communities. 2013. doi:10.48550/ARXIV.1301.3120

29. Li Z, Zhang S, Wang R-S, Zhang X-S, Chen L. Quantitative function for community detection. Phys Rev E Stat Nonlin Soft Matter Phys. 2008;77: 036109.

30. Newman MEJ, Girvan M. Finding and evaluating community structure in networks. Phys Rev E Stat Nonlin Soft Matter Phys. 2004;69: 026113.

31. Aldecoa R, Marín I. Surprise maximization reveals the community structure of complex networks. Sci Rep. 2013;3: 1060.

32. Coleman J. Relational analysis: The study of social organizations with survey methods. Hum Organ. 1958;17: 28–36.

33. Donker T, Smieszek T, Henderson KL, Johnson AP, Walker AS, Robotham JV. Measuring distance through dense weighted networks: The case of hospital-associated pathogens. PLoS Comput Biol. 2017;13: e1005622.

34. Diekmann O, Heesterbeek JAP. Mathematical Epidemiology of Infectious Diseases: Model Building, Analysis and Interpretation. John Wiley & Sons; 2000.

35. Anderson RM, May RM. Directly transmitted infections diseases: control by vaccination. Science. 1982;215: 1053–1060.

36. van Rossum G. Python tutorial. 1995 Jan. Report No.: R 9526. Available: https://ir.cwi.nl/pub/5007/05007D.pdf

37. Hagberg A, Swart P, S Chult D. Exploring network structure, dynamics, and function using networkx. Los Alamos National Lab. (LANL), Los Alamos, NM (United States); 2008 Jan. Report No.: LA-UR-08-05495; LA-UR-08-5495. Available: https://www.osti.gov/servlets/purl/960616

38. Rossetti G, Milli L, Cazabet R. CDLIB: a python library to extract, compare and evaluate communities from complex networks. Appl Netw Sci. 2019;4. doi:10.1007/s41109-019-0165-9

39. Oliphant TE. Python for Scientific Computing. Comput Sci Eng. 2007;9: 10–20.

