## Supplementary Information for "Anticipating the risk and spatial spread of measles in populations with high MMR uptake: using school-household networks to understand the 2013 - 2014 outbreak in the Netherlands"

**Supplementary information: School-Household networks, untangling the complexities of measles epidemiology in the Netherlands**

*Evaluation of the resolution parameter*

We created partitions using the leiden algorithm varying the resolution parameter between 0.1 and 1.0. We tested the partitions using three metrics:

*Internal edge density*: This metric measures the proportion of possible edges that are present in community C, expressed as:

$$\rho_C = \frac{m_C}{½\, n_C(n_C - 1)}$$

where $m_C$ is the number of edges internal to community $C$ and $n_C$ is the number of schools in community C. This takes a value between 0 and 1 and provides a quantification of the absolute connectivity within the community, irrespective of connectivity with other communities.

*Modularity density*: Modularity density normalises the quality function by the number of schools in the community, hence removing dependence on community size. This provides a better comparison of modularity between partitions with different community sizes. This is expressed:

$$Q_{dens}(S) = \sum_{C\in S}\frac{1}{n_C}\left(\sum_{i\in C}k_{iC}^{in} - \sum_{i\in C}k_{iC}^{out}\right)$$

where $n_C$ is the number of schools in $C$, $k_{iC}^{in}$ is the degree of node i within $C$ (edges to schools inside the community) and $k_{iC}^{in}$. is the degree of node 8 outside P (edges to schools outside the community) for a partition $S$.

*Newman Girvan Modularity:* A classic metric of how strongly defined communities in a partition are from the rest of the graph. Is calculated as:

$$Q_{NG}(S) = \frac{1}{m}\sum_{C \in S}\left(m_C - \frac{(2m_C + l_C)^2}{4m}\right)$$

where $m$ is the number of graph edges, $m_C$ is the number of community's edges, $l_C$ is the number of edges from schools in $C$ to schools outside $C$.

*Surprise:* Surprise is a quality metric assuming that edges between nodes emerge according to a hyper-geometric distribution. According to the Surprise metric, the higher the score, the less likely that the communities detected occurred at random and therefore the better the quality of the partition.

We found that all metrics were optimised when the resolution parameter was set to 1.0 (Figure S1), which results in the “unmodified” Leiden algorithm.

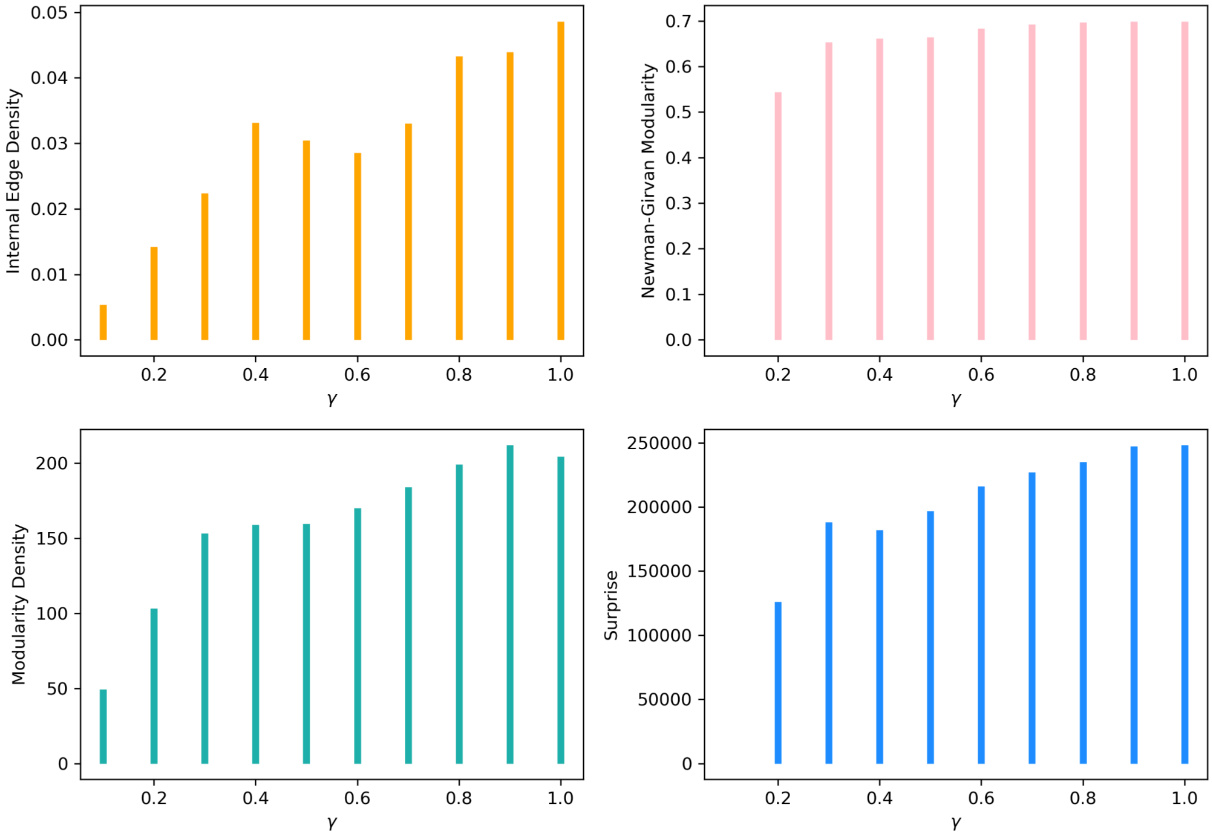


**Figure S1**. Quality metrics for various values of resolution parameter + for partitions recovered using the modified Leiden algorithm. Panels A to D show the scores for Internal Edge Density, Newman-Girvan Modularity, Modularity Density and Surprise respectively

*Translation of contact network to a school based transmission model*

The basis of the model is that the network of contact between schools can provide an estimate of the probability that individual school outbreaks can seed an outbreak in each neighbouring school.

For each pair of neighbouring schools, we calculated the probability that an outbreak could be seeded in school $i$ given that an outbreak does occur in adjacent school $j$. First we consider the probability of transmission between siblings, in the event that one is infectious and the other is susceptible, to be a set value $q$. The probability that the child from school $j$ is infected is denoted by $P_j^I$ and the probability that the child from school j is susceptible by $P_i^S$. The probability that a single infected student in school *i* causes a large outbreak in that school is $P_i^{OB}$. The probability of an outbreak in school $j$ leading to an outbreak in school $i$ through each unique contact pair that link schools $i$ and $j$ is:

$$P_j^I P_i^S q P_i^{OB}$$

The probability that the child in school $j$ is infected, $P_j^I$, was assumed to be equal to the proportion of the school children infected by the outbreak in that school. we assumed that this is defined by the solution of the final size equation[19]:

$$R_j(\infty) = \left(1 - V_j\right) e^{-\left(1 - V_j\right) R_0 R(\infty)}$$

Where $V_j$ is the vaccination coverage in school $j$.

The probability that the child in school $i$ is susceptible is equal to the proportion of school i that remains unvaccinated, $(1 - V_i)$.

I took the probability of an outbreak occurring in that school as a result of this transmission to be:

$$P_i^{OB} = \left(1 - \frac{1}{R_{eff}}\right) = \left(1 - \frac{1}{\left(1 - V_i\right) R_0}\right)$$

Which assumes a Poisson distribution of within school contact rate amongst children[19].

The probability that none of the unique contact pairs causes an outbreak in school i can be written:

$$\prod_{All\,pairs}\left(1 - P_j^I P_i^S q P_i^{OB}\right) = \left(1 - P_j^I P_i^S q P_i^{OB}\right)^{C_{ij}}$$

Therefore, the probability that at least one contact pair causes an outbreak in school $i$ is the complement of this:

$$P_{trans,\ ij} = \left[1 - \left(1 - P_j^I P_i^S q P_i^{OB}\right)^{C_{ij}}\right]$$

This provides a basis upon which to model simulations of outbreaks across networks of schools in different settings.

The simulations in the analysis detailed in this paper were performed by generating A binary decision network of transmission, where edges were weighted 1 or 0 to indicate transmission or no transmission between schools. The edges of the binary network were set by drawing from a binomial distribution with probability of transmission calculated as detailed above. The extent of the outbreak was defined as the sum of the out components of initially infected schools.

*Table S1 school denominations*

| School Denomination | *Dutch name* | *Schools* | *Primary* | *Secondary* |
|---|---|---|---|---|
| Public school | Openbaar | 2810 | 2466 | 344 |
| Roman Catholic | Rooms-Katholiek | 2554 | 2258 | 296 |

| | | | | |
|---|---|---|---|---|
| Mainstream protestant | Protestants-Christelijk | 2147 | 1904 | 243 |
| Special educational philosophy | Algemeen bijzonder | 807 | 579 | 228 |
| Dutch Reformed | Reformatorisch | 208 | 181 | 27 |
| Reformed liberated | Gereformeerd vrijgemaakt | 118 | 118 | 0 |
| Anthroposophic | Antroposofisch | 81 | 70 | 11 |
| Islamic | Islamitisch | 44 | 43 | 1 |
| Interconfessional | Interconfessioneel | 21 | 15 | 6 |
| Reformed Liberated | Gereformeerd | 18 | 0 | 18 |
| Evangelical | Evangelisch | 16 | 12 | 4 |
| Hindu | Hindoeistisch | 6 | 6 | 0 |
| Other | Overige | 4 | 0 | 4 |
| Jewish | Joods | 2 | 2 | 0 |
| Moravian Church | Evangelische Broedergemeenscha | 2 | 2 | 0 |
| Potestiant/Evangelical | Protestants-Christelijk/Evange | 1 | 0 | 1 |
| Jewish Orthodox | Joods orthodox | 1 | 0 | 1 |
| Potestiant/Reformed | Protestants-Christelijk/Reform | 1 | 0 | 1 |

*Alternative model 2: Spatial interaction between schools*

The spatial interaction model was designed to have a broadly equivalent spatial distribution of contact between schools but with otherwise naïve interaction (i.e. no preference for contact within particular denominations or between tiers of education etc.).

We formulated interaction between schools as an exponentially distributed distance relation with an additional weighting on schools determined by their degree in the empirical school network. We selected the parameterisation of this distribution by visually matching the resultant distribution of the *distance between schools connected by contact pairs* over the entire network. The value of $\lambda$ that best reflected the data-driven network was 0.65, which was aimed at matching the peak of the distribution of contact described in the data.

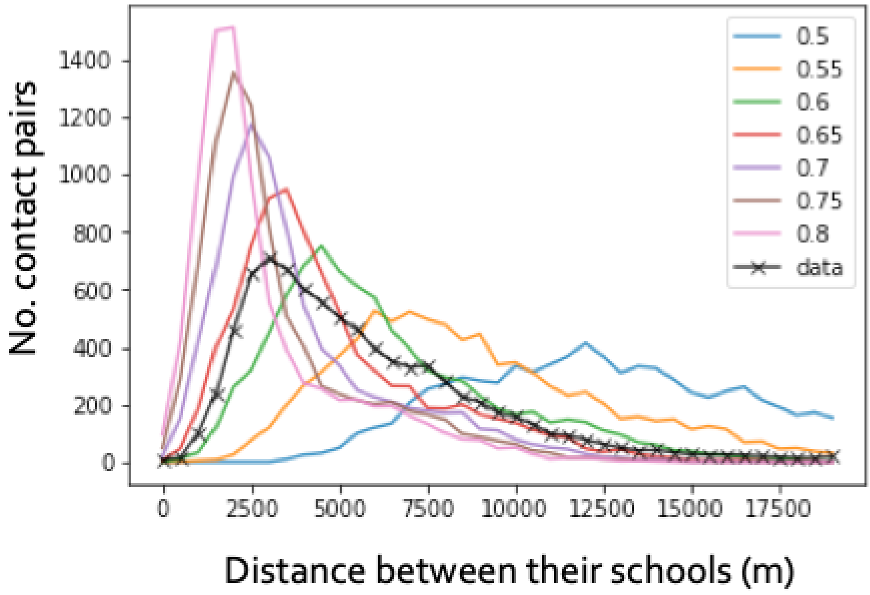


**Figure S2** The distribution of distance between schools connected by contact pairs. The coloured lines show interaction based on spatial interaction models with parameters between 0.5 and 0.8. black line with cross markers shows the distribution in the household links from the data.

*Outbreak simulations*

The binary outbreak networks were used to identify schools, which would be infected by an outbreak initiated in each school in the network. This was achieved using the following approach:

The binary outbreak network is a directional graph with edges weighted either 1 or 0. 1 means transmission occurs between the schools in the direction of the edge.

To calculate the risk posed by a school, the schools eventually infected are those connected by chains of out-edges (schools infected by the previous generation of the outbreak) on the binary outbreak network. Figure S3 shows schools in progressive generations of an outbreak on an example network. The school of interest (i.e. the school posing the risk) is coloured red. Each network shows a different generation of the outbreak, where newly infected schools are shown in green and previous generations are shown in blue. There are 4 generations of transmission after the initial school infected and10 schools are eventually infected by the outbreak.

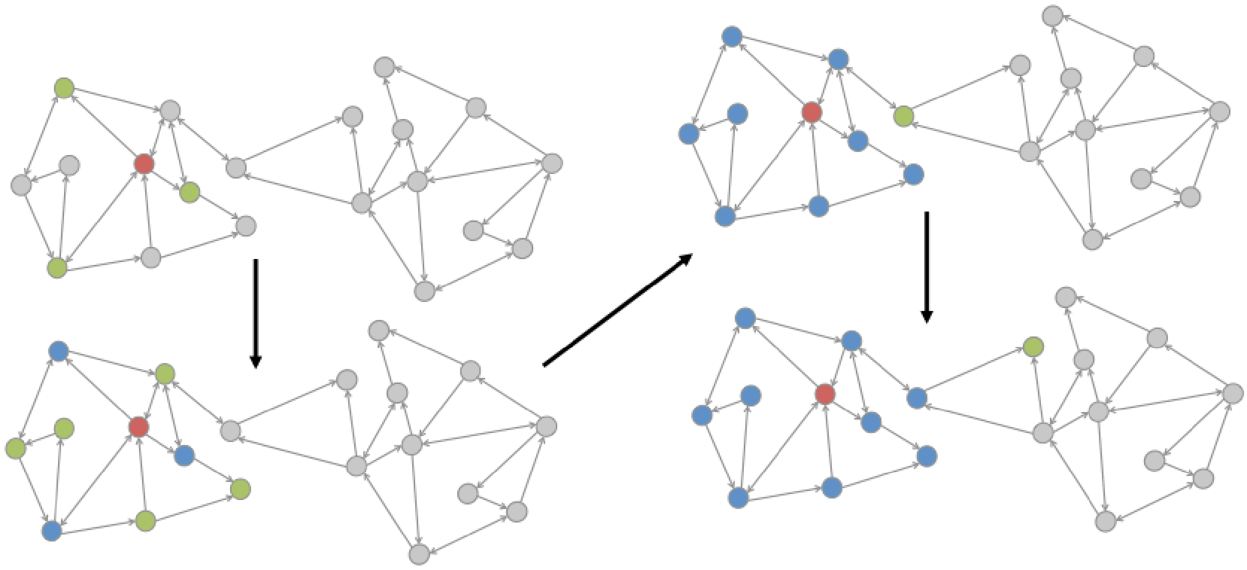

**Figure S3** Schematic of a network showing chains of out-edges to form the out component of a node as a method for finding the schools at risk of infection from an outbreak initiated in a particular school. The initial school is shown in red, each new generation of schools connected by out-edges is shown in green, previous generations are shown in blue. Black arrows indicate the direction of successive generations

Supplementary figures

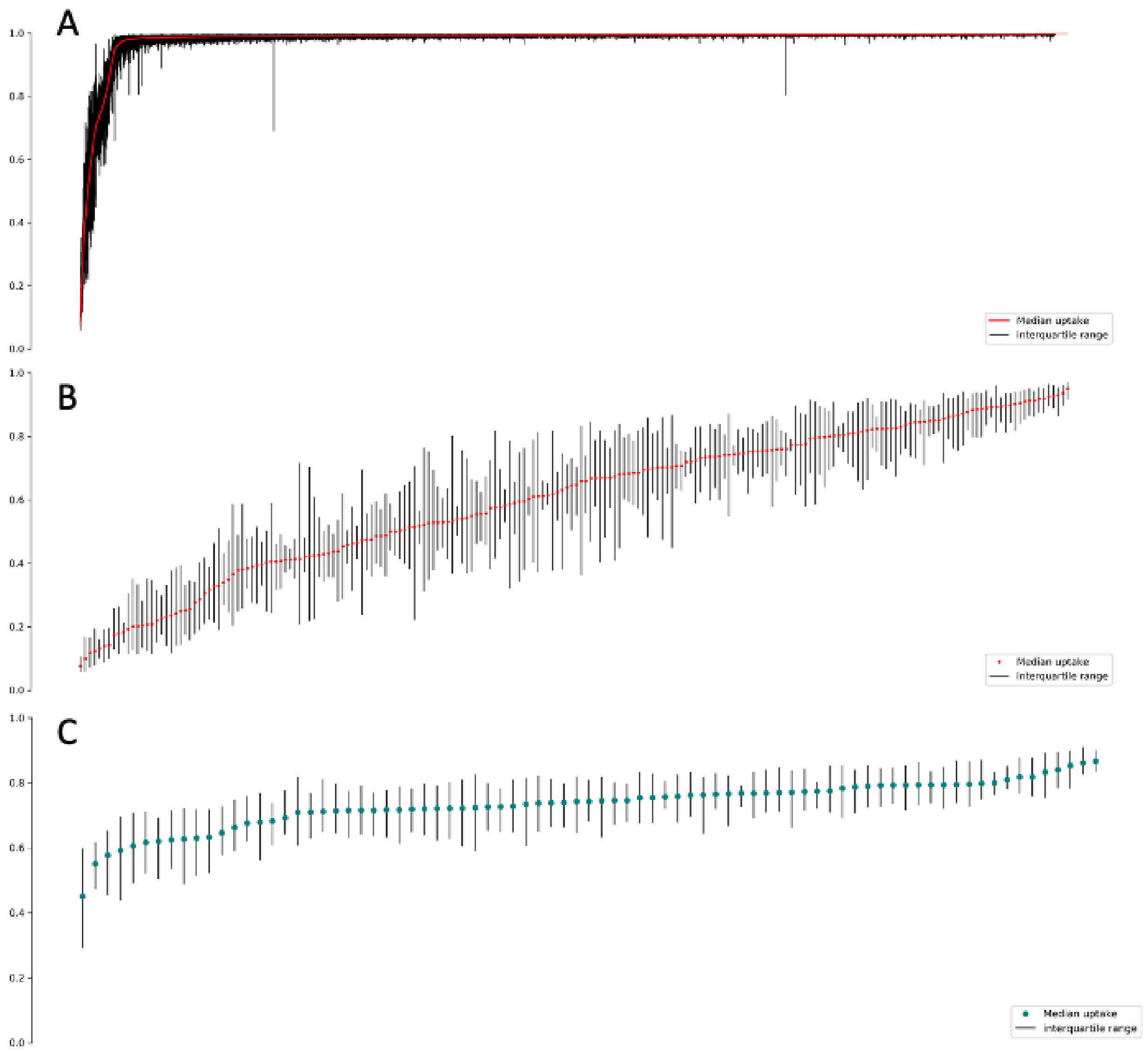


**Figure S4** estimates of vaccine uptake by school, showing median values and interquartile range for each school estimated by Klinkenberg et al. for A) all schools, B) Orthodox Protestant schools and C) Anthroposophic schools.
